# Sleep Apnoea and Memory (SAM): protocol for a prospective study of prevalence and symptoms of sleep apnoea in memory clinics

**DOI:** 10.64898/2026.03.09.26347943

**Authors:** Victoria Grace Gabb, Charlotte Neary, Devan Mair, Adrian Kendrick, Gregor Russell, Julie Clayton, Samina Begum, Robert Huckstepp, Nicholas Turner, Elizabeth Coulthard

## Abstract

**Introduction:** Sleep apnoea is common in older adults and a risk factor for cognitive decline and dementia but is rarely assessed in memory clinics. The Sleep Apnoea and Memory (SAM) study will assess the prevalence of sleep apnoea and identify optimal screening for sleep apnoea in memory clinics.

**Methods:** SAM is a prospective observational multi-site study recruiting adults attending NHS memory clinics. Participants will undergo a single night of polygraphy using a home sleep apnoea test (WatchPAT™ 300) and complete questionnaires based on NICE guidance for sleep apnoea assessment. The primary outcome will be the prevalence of sleep apnoea. Secondary outcomes include determining sleep apnoea prevalence across different cognitive diagnoses, identifying which symptoms and risk factors which best predict sleep apnoea, and assessing feasibility of remote sleep apnoea screening.

**Discussion:** The SAM study will improve understanding of the extent of sleep apnoea in people attending memory clinics and inform design of an interventional trial for treating sleep apnoea in patients with cognitive impairment. Treating sleep apnoea in memory clinics may help to improve symptoms and/or prognosis for people experiencing memory problems.

## 1 Introduction

Sleep apnoea is a common sleep-related breathing disorder, particularly in older adults, in which breathing is frequently interrupted during sleep through complete pauses in breathing (apnoeas) and periods of slower or more shallow breathing (hypopneas) (Ghavami et al., 2023). Patients with sleep apnoea may present with daytime symptoms, such as daytime sleepiness or headaches, nighttime symptoms such as disturbed sleep and snoring, or may report no symptoms (Lacasse et al., 2002).

Sleep apnoea can broadly be categorised into obstructive sleep apnoea (OSA) and central sleep apnoea (CSA), though patients may present with both types (Zhang et al., 2023). OSA, where there is respiratory effort but no or partial airflow due to an upper airway blockage, is the most common type of sleep apnoea (Benjafield et al., 2019, Jordan et al., 2014). CSA, where both airflow and respiratory effort are reduced, is less common, typically resulting from comorbidities such as heart failure or stroke, or a side effect of medications (Randerath et al., 2024). Pauses in respiration result in recurrent intermittent hypoxia and hypercapnia, sleep fragmentation, increased sympathetic activity, and oxidative stress (Henning and Anderson, 2025, Bahia and Pereira, 2015). It is therefore unsurprising that sleep apnoea is associated with increased risk of cardiovascular disease, stroke, metabolic dysfunction, accidents, and mortality (Knauert et al., 2015). Despite advances in diagnosis and treatment of sleep apnoea, a high proportion of cases remain undiagnosed or ineffectively managed (Iannella et al., 2025).

The prevalence of sleep apnoea increases with age, with 1 in 7 adults (Benjafield et al., 2019) and 1 in 3 older adults (Ghavami et al., 2023) estimated to have sleep apnoea. Ageing is associated with various changes to sleep, which could trigger or exacerbate sleep apnoea. During aging, sleep becomes more fragmented, and increased arousal frequency can destabilise breathing, whilst changes to the upper airway, such as lengthening of the pharyngeal airway and reduced response to negative pressure, can predispose it to collapse (Glasser et al.). Whilst sleep apnoea is more prevalent in men than premenopausal women, the prevalence of sleep apnoea in women increases markedly during and after menopause, likely influenced by changes to sex hormones and body composition (Jehan et al., 2016) (Mirer et al., 2017). Additionally, older adults have a higher risk of multiple comorbidities hypothesised to have a bidirectional relationship with sleep apnoea, including diabetes (Muraki et al., 2018) and cardiovascular disease (Yeghiazarians et al., 2021, Glasser et al.).

Sleep apnoea is also increasingly recognised as a potentially modifiable risk factor for cognitive decline and dementia (Lajoie et al., 2020, Gurbani et al., 2025, Ercolano et al., 2024). Untreated sleep apnoea has been associated with deficits in memory, attention, and executive function (BUCKS et al., 2013) and patients with symptomatic OSA are more likely to develop mild cognitive impairment (MCI) or Alzheimer’s disease (AD) (Bubu et al., 2020, Tsai et al., 2020). Elevated levels of several AD fluid biomarkers, including amyloid-beta (Aβ) and tau pathology, oxidative stress, and inflammation are associated with sleep apnoea (Baril et al., 2018). Treating sleep apnoea can improve sleep and quality of life both in patients and their bed partners (Siccoli et al., 2008, Labarca et al., 2020, Fietze et al., 2023, Luyster, 2017), is associated with lower cerebrospinal fluid (CSF) Aβ burden (Ju et al., 2019), and improve cognitive outcomes (Benkirane et al., 2024, Costa et al., 2023). It may have socioeconomic benefits through reducing healthcare utilisation costs and risk of accidents and improving productivity (Watson, 2016, Zappalà et al., 2025). Effective treatments for sleep apnoea include positive airway pressure (PAP), oral appliances, weight loss, hypoglossal nerve stimulation, and surgery to enlarge the upper airway (Gottlieb and Punjabi, 2020), though continuous positive airway pressure (CPAP) is often considered the first line treatment.

Identifying and treating sleep apnoea in memory clinics may therefore represent an opportunity to improve quality of life, and cognitive and health outcomes. However, it is not yet understood how prevalent sleep apnoea is in patients presenting with cognitive symptoms, as sleep is rarely assessed in memory clinics. One recent study conducted at a memory clinic in Australia identified OSA in 75% of their memory clinic patients, with only 13% previously having a diagnosis. In this sample, patients with OSA also had worse global cognition and memory performance compared to memory clinic patients without OSA (Lam et al., 2025b). In recent years, there has been increasing recognition that screening for sleep apnoea in patients presenting with cognitive impairment is important (Liguori, 2025). For example, both the Canadian Consensus Conferences on the Diagnosis and Treatment of Dementia (Ismail et al., 2020) and Sleep Study Group of the Italian Dementia Research Association (Guarnieri et al., 2014) recommend that patients at risk for dementia should be screened for symptoms of sleep apnoea.

Validated questionnaires such as the STOP-Bang (Chung et al., 2016) (designed to assess risk of sleep apnoea) and Epworth Sleepiness Scale (Johns, 1991) (ESS; designed to assess daytime sleepiness) are often used to identify individuals at high risk of sleep apnoea who should undergo oximetry or polygraphy including in primary care and perioperative settings (Rosenthal and Dolan, 2008, Senaratna et al., 2019). However, their predictive performance is limited, particularly in older adults and memory clinic populations (Lam et al., 2025a, Martins et al., 2020, Jorge et al., 2019). Older adults typically present with fewer sleep apnoea symptoms compared to younger adults, and the commonest symptoms in older adults (such as sleepiness, memory loss, nocturia, and sexual dysfunction) may be attributed to ageing and/or neurodegeneration (Antonaglia et al., 2025). Therefore, objective data collection and identifying the most appropriate questionnaires for older adults and individuals with cognitive impairment is warranted.

In adult populations, including older adults, in-laboratory polysomnography (PSG) is considered the gold standard (or Type I) test for sleep apnoea diagnosis in older adults. This collects data across many channels including electroencephalogram (EEG), electrooculogram, electromyogram, and an electrocardiogram (ECG) (**Table 1**). However, PSG requires significant resources and can be burdensome for patients, meaning it is impractical to use at scale. Home sleep apnoea tests (HSATs) or Type III respiratory polygraphy devices have the potential to improve access to sleep apnoea diagnosis at scale, without requiring a visit to a sleep clinic or other healthcare provider (Riha et al., 2023). Several HSATs have been approved for use by the Food and Drug Administration (FDA) and National Institute for Health and Care Excellence (NICE) in diagnosing sleep apnoea, including the WatchPAT® and AcuPebble™(Park et al., 2024, NICE, 2024).

**Table 1.**
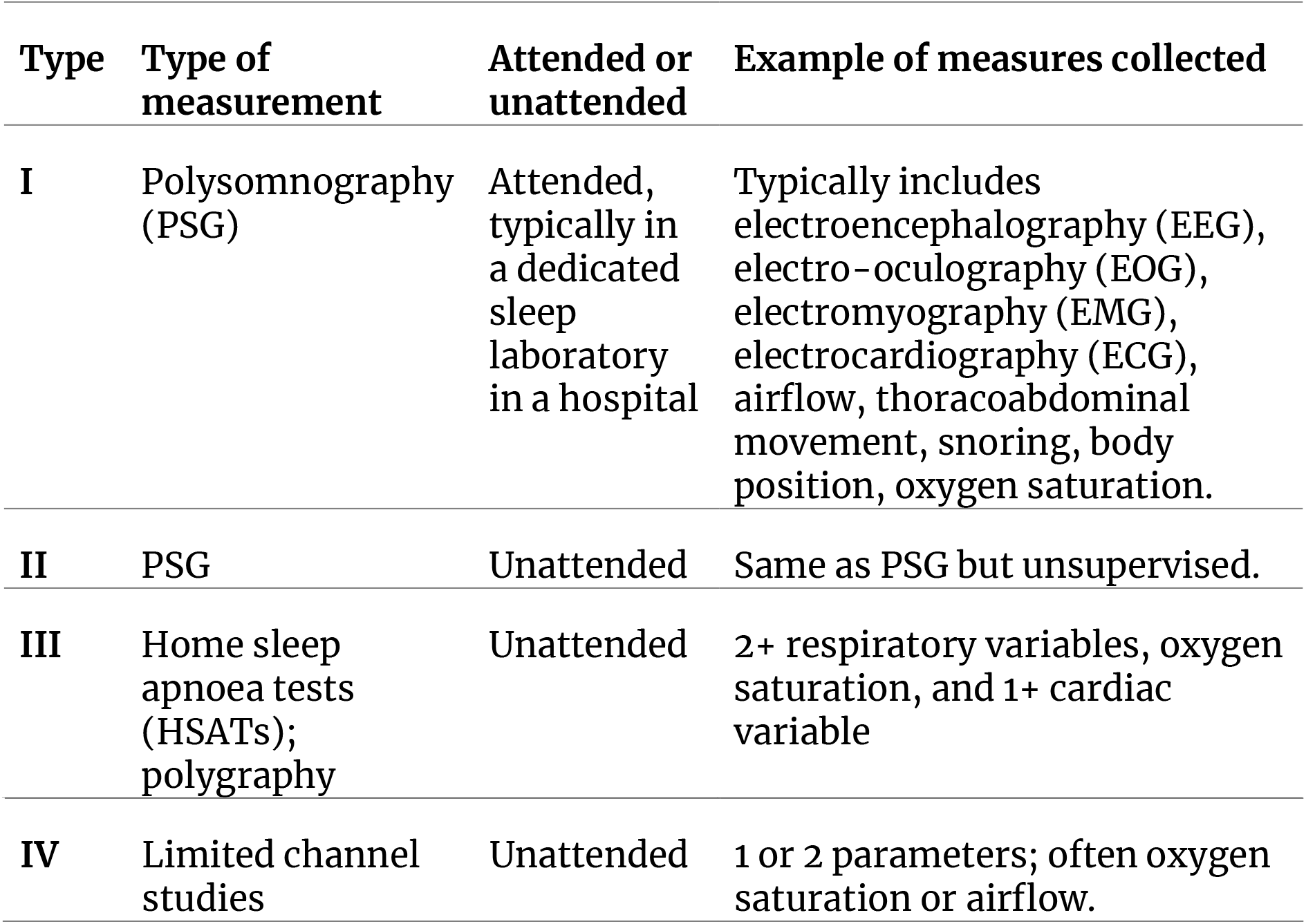
Sleep studies used in sleep apnoea screening and diagnosis: type I, II, III, and IV.

| Type | Type of measurement | Attended or unattended | Example of measures collected |
| --- | --- | --- | --- |
| I | Polysomnography (PSG) | Attended, typically in a dedicated sleep laboratory in a hospital | Typically includes electroencephalography (EEG), electro-oculography (EOG), electromyography (EMG), electrocardiography (ECG), airflow, thoracoabdominal movement, snoring, body position, oxygen saturation. |
| II | PSG | Unattended | Same as PSG but unsupervised. |
| III | Home sleep apnoea tests (HSATs); polygraphy | Unattended | 2+ respiratory variables, oxygen saturation, and 1+ cardiac variable |
| IV | Limited channel studies | Unattended | 1 or 2 parameters; often oxygen saturation or airflow. |

### Study objectives

Our broad objective is to work towards future clinical trials treating sleep apnoea in memory clinics to see if treatment slows progression and/or improves patients’ quality of life. The present study aims to find out the prevalence of sleep apnoea, the feasibility of remote sleep apnoea testing, and whether sleep apnoea treatment may improve blood biomarkers relating to Alzheimer’s disease and neurodegeneration in patients attending memory clinics.

Specifically, the study will:

1. Determine the proportion of people attending memory clinics who have at least mild sleep apnoea, as defined by an oxygen desaturation index (ODI) 3% of ≥ 5, and whether prevalence differs by cognitive diagnosis.
2. Identify the symptoms or risk factors that best predict sleep apnoea in people who present to memory clinics.
3. Assess the feasibility of remote questionnaires for sleep apnoea screening in memory clinics.
4. Pilot methodology for observing the effect of treating sleep apnoea on blood biomarkers of Alzheimer’s disease and neurodegeneration.

## 2 Methods and Analysis

This is a pragmatic, prospective, observational, multi-site study designed to assess the prevalence and predictors of sleep apnoea in memory clinic patients. The study is sponsored by North Bristol NHS Trust and funded by the National Institute of Health and Care Research (NIHR) Research for Patient Benefit programme (NIHR207185). The study was prospectively registered on the International Standardized Randomized Controlled Trial Number (ISRCTN) registry (reference: ISRCTN11309942). The expected recruitment period is 10 months from the first site opening.

### Ethics and informed consent

The study received a favourable opinion from the Health Research Authority and North West - Greater Manchester South Research Ethics (reference: 25/NW/0221). The study will be explained by a member of the research team, and each prospective participant will be provided with a participant information sheet to explain the purpose of the study, the study procedures, and implications for the patient if they are found to experience daytime sleepiness or are diagnosed with sleep apnoea. Patients will be given time to consider whether they would like to participate and the opportunity to ask questions and discuss their involvement with others, such as partners or relatives, before being asked to consent. Written informed consent will be obtained from all participants prior to engaging in study procedures.

### Participants

#### Recruitment

Eligible participants will be adults aged 18 or over attending memory or cognitive disorders services at participating NHS sites (in Bristol, London, Cornwall, Northumbria, and Bradford) or affiliated participant identification centres in the six months prior to the date of consent. Patients may be approached prior to, or at, their clinical appointment, or within six months of an appointment at a participating site. Appointments may be initial consultations or follow-up appointments, and patients will not be excluded based on any diagnosis. As the primary aim is estimating sleep apnoea prevalence, individuals with an existing diagnosis of sleep apnoea will also be eligible for inclusion and will be asked to continue their usual treatment for the duration of the study.

#### Sample size

A sample size of 385 participants was chosen to allow for estimation of 50% prevalence and investigation of 16 parameters as predictors for sleep apnoea, and balances accuracy with model overfitting. To allow for a 15% drop-out, the overall recruitment target is 453 participants with data from 385 patients. Recruitment targets will vary across sites based on research capacity and the number of eligible patients.

### Study setting

The study is designed to be delivered fully remotely, using a home sleep apnoea test (WatchPAT™ 300) and an electronic data capture system (REDCap) for consent and data collection. However, some study procedures may also take place over the telephone or face-to-face (e.g., at the memory clinic or in a participant’s home) to minimise digital exclusion (Wilson-Menzfeld et al., 2025).

### Study procedures

Participants will undergo study assessments as shown in **Table 2**.

**Table 2.**
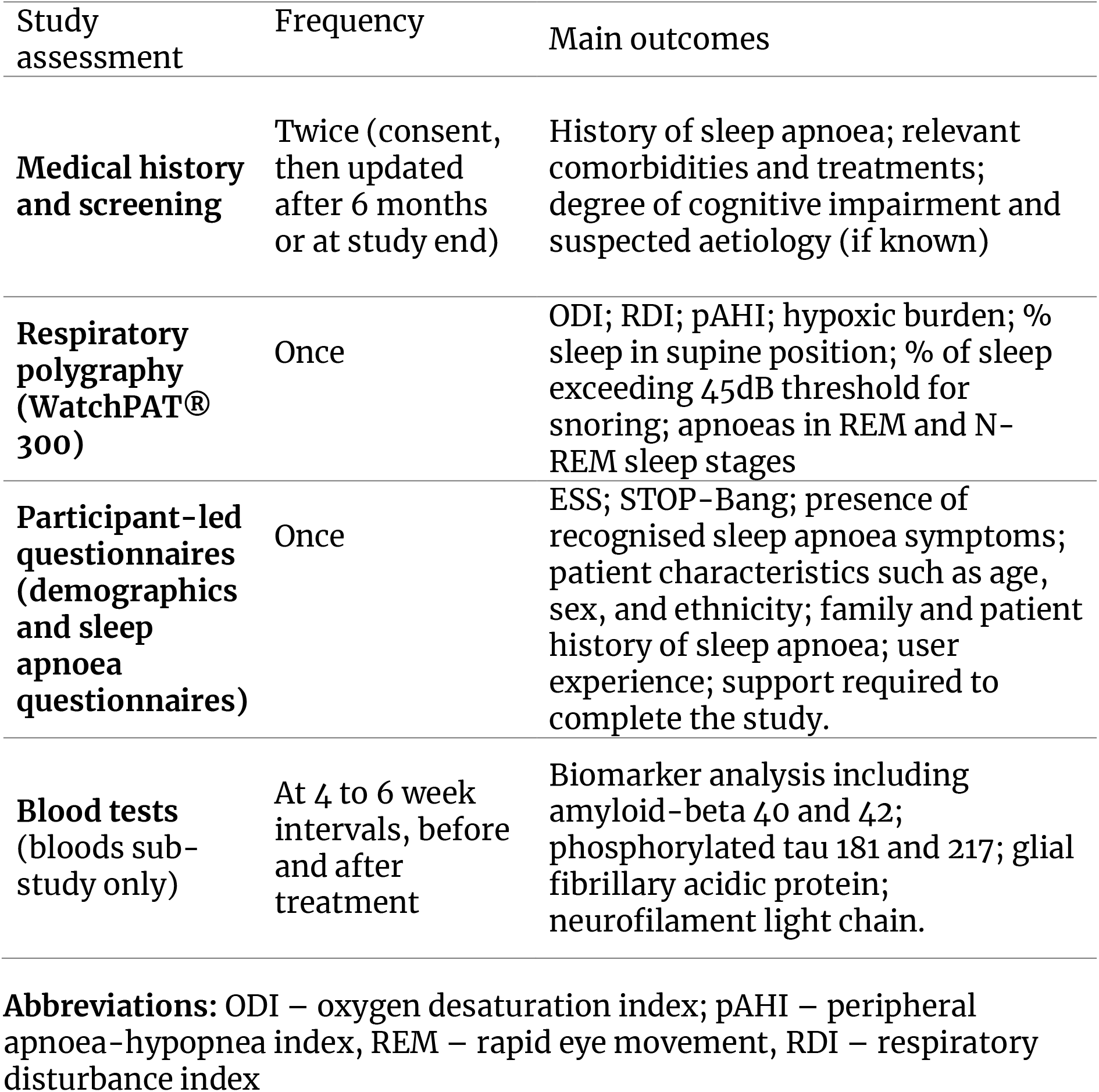
Summary of study assessments for the Sleep Apnoea and Memory (SAM) study.

#### Main study

##### Screening and Eligibility

Eligibility will be confirmed by a member of the research team. Data on relevant diagnoses, medications, and medical history will be obtained from medical records and/or directly from patients, including information on the most recent cognitive diagnosis and scores from recent cognitive assessments. Due to the likelihood of new diagnoses or cognitive changes, where available, medical records will be reviewed at 6 months and at the end of the study for any relevant cognitive updates (e.g., new diagnosis, biomarker results).

##### Home Sleep Apnoea Testing

Participants will be asked to complete a single night of respiratory polygraphy using the WatchPAT® 300, a HSAT device recommended by NICE (NG202) (NICE, 2021). The WatchPAT® 300 collects data across seven channels from three points of contact (at the fingertip, chest, and wrist): peripheral arterial tone (PAT) signal, heart rate, oximetry, actigraphy, body position, snoring, and chest motion. Earlier versions (WatchPAT® 100 and 200) have been demonstrated to have good sensitivity and acceptable specificity compared to conventional PSG (Moffa et al., 2023) and have been previously used in patients with MCI and AD dementia (Tadokoro et al., 2020).

Participants will also be asked to complete a demographics questionnaire and a questionnaire for symptoms and risk factors of sleep apnoea, including the ESS (Johns, 1991) and STOP-Bang (Chung et al., 2016) alongside bespoke items as recommended by NICE (NICE, 2021). The questionnaire will also include more general questions around sleep disturbance and include single screening questions for restless legs syndrome and rapid eye movement (REM) sleep behaviour disorder (Ferri et al., 2007, Postuma et al., 2012). We will also ask patients to rate how easy or difficult the device was to use, as well as provide general feedback on their experience (Riha et al., 2023).

The WatchPAT® 300 devices will be provided to participants via the dedicated WatchPAT® Direct service, who will also provide participants with written instructions for use and pre-paid envelopes for return postage (or arrange home collection where necessary). Upon return, the WatchPAT® Direct team will upload the data to a secure cloud-based platform, *CloudPAT®*. Data from the WatchPAT® 300 will be manually reviewed alongside data collected from the medical history and patient-led questionnaires by a sleep physiologist with experience in interpreting polygraphy for sleep apnoea assessments.

The participant and their GP will be informed of the study results by the local site team. Standard local procedures will be followed if referrals for further investigation, treatment, or follow-up are indicated.

#### Bloods sub-study

Participants at the lead participating site (North Bristol NHS Trust) who receive a new diagnosis of sleep apnoea from the main study and are referred for treatment will be invited to participate in a blood biomarker sub-study.

This sub-study aims to observe whether there are changes in blood biomarkers relevant to neurodegeneration and AD after sleep apnoea treatment. Participants will be asked to undergo a series of blood tests approximately 4 to 8 weeks apart, spanning the period leading up to and after their treatment has started. Blood plasma will undergo biomarker analysis, likely including the Aβ42/40 ratio, phosphorylated tau (p-tau) 181, p-tau 217, glial fibrillary acidic protein, and neurofilament light chain. However, we may also integrate emerging candidate biomarkers relating to AD or neurodegeneration (Grigoli et al., 2024). This will give us an indication if treating sleep apnoea may slow the progression of AD.

### Main outcomes

The primary outcome will be the proportion of people in memory clinics who have at least mild sleep apnoea (ODI 3% ≥ 5). Secondary outcomes will include: the proportion of people with mild sleep apnoea with different aetiological diagnoses (e.g., Alzheimer’s disease, Lewy body disease); the proportion of people with mild, moderate (ODI 3% ≥15<30), and severe (ODI 3% ≥ 30) sleep apnoea; proportion of mild sleep apnoea with more conservative cut-off of ODI 4% > 5); diagnostic accuracy of the STOP-Bang, ESS, and additional questions for at least mild sleep apnoea; data completeness for questionnaires completed online, via telephone, or face- to-face; and change in blood biomarkers over time.

### Patient and public involvement

Patient and public involvement (PPI) representatives with lived experience of cognitive impairment and/or sleep apnoea have been involved since study inception. SB, who has lived experience as a carer for someone living with dementia, is part of the research team, having supported the initial funding and ethics applications, and providing continued oversight of the study to ensure public perspectives are included throughout the full research process. Additional PPI representatives have advised on the relevance of the project and helped to develop the study protocol and patient-facing materials, including advising on wording for the bespoke questions in the sleep apnoea questionnaire. PPI representatives will also be invited to regular meetings with the research team and steering committee.

### Data analysis

Descriptive statistics will summarise participant demographics,clinical characteristics, sleep questionnaire responses (e.g., STOP-Bang, ESS), and objective sleep parameters derived from the WatchPAT.

For sleep apnoea prevalence outcomes, descriptive statistics will be provided alongside 95% confidence intervals. Prevalence will be determined where the number of cases for a given aetiological diagnosis exceeds ten participants. Primary analyses will be calculated at 3% ODI cut-offs with sensitivity analyses of 4% ODI cut-offs and for 3% and 4% AHI cut-offs. We will also report the proportion of patients recommended for referral for treatment or further investigation due to sleep apnoea and other incidental findings (e.g., possible periodic limb movements or atrial fibrillation).

For predictors of sleep apnoea, area under the ROC curve will be calculated alongside sensitivity and specificity to individually assess accuracy of the ESS and STOP-Bang as well as individual component scores. Adaptive least absolute shrinkage and selection operator (LASSO) will be used to explore predictive models for sleep apnoea. The proportion of data completeness will be reported for each predictor and outcome variable along with the proportion of complete cases and where available, reasons for missing data. Multiple imputation may be used in prediction model development.

Participant flow will be reported in line with STROBE guidance (von Elm et al., 2007). We will record the number of people who opt to complete the questionnaires online, via telephone, and face-to-face with site staff and data completeness for each method.

Interrupted time series analysis will be used, with a time series model fit with linear regression using each biomarker as an outcome including time as a predictor, an indicator variable to represent pre-intervention or postintervention period, and a time by intervention period indicator variable interaction.

## 3 Discussion

This project will support development of future clinical trials testing the effectiveness of sleep apnoea treatment in people with cognitive impairment and neurodegeneration. Currently, treatments for patients attending memory clinics are limited. Sleep, and sleep apnoea specifically, can be improved with interventions, and may represent a modifiable risk factor for dementia. Identifying the prevalence and symptoms of sleep apnoea in memory clinics, and the feasibility of remote testing, are important steps towards future clinical trials, where we may be able to slow disease progression and/or improve cognitive outcomes and quality of life.

## 4 Conflict of Interest

The authors declare that the research was conducted in the absence of any commercial or financial relationships that could be construed as a potential conflict of interest.

## 5 Author Contributions

**VGG:** Conceptualisation, Methodology, Resources, Data Curation, Writing – Original Draft, Writing – Review & editing, Visualisation, Formal analysis, Supervision, Project administration, Funding acquisition. **CN:** Writing – Original Draft, Writing – Review & editing, Resources, Investigation. **DM:** Writing – Original Draft, Writing – Review & editing, Investigation. **AK:** Conceptualisation, Methodology, Formal analysis, Writing – Review & editing, Supervision, Funding acquisition. **GR:** Conceptualisation, Supervision, Investigation, Writing – Review & editing. **JC:** Conceptualisation, Methodology, Funding acquisition, Writing – Review & editing. **SB:** Conceptualisation, Methodology, Funding acquisition, Writing – Review & editing. **RH:** Supervision, Writing – Review & editing. **NT:** Conceptualisation, Methodology, Formal analysis, Writing – Review & editing. **EC:** Conceptualisation, Methodology, Investigation, Resources, Writing – Review & editing, Supervision, Project administration, Funding acquisition.

## 6 Funding

This study is funded by the National Institute of Health and Care Research (NIHR) Research for Patient Benefit programme (NIHR207185). The views expressed are those of the authors and not necessarily those of the NIHR or the Department of Health and Social Care. The study is also supported by a philanthropic donation from the Goldman Foundation.

## 7 Acknowledgments

The authors would like to thank the lived experience experts who supported the grant application and study development.

## 9 Data Availability Statement

The datasets generated for this study will be made available in the University of Bristol’s publicly accessible Research Data Repository data.bris, https://www.bristol.ac.uk/staff/researchers/data/accessing-research-data/.

## Notes

### Competing Interest Statement

The authors have declared no competing interest.

### Author Declarations

The Health Research Authority and North West - Greater Manchester South Research Ethics Committee gave ethical approval for this work (reference: 25/NW/0221).

